# Direct Percutaneous Embolization of Head, Neck and Spine Tumors: A Single Center Experience

**DOI:** 10.1101/2024.07.09.24310047

**Authors:** Karan Topiwala, Shiwei Huang, Luke Sabal, Bailey Kahmeyer, Andrew Grande, Bharathi Jagadeesan

## Abstract

**Introduction:** Transarterial embolization is a well-established adjunct in the management of hypervascular head, neck, and spine tumors. Few case-series have described the role of direct percutaneous embolization (DPE).

**Method:** We report our experience with DPE (with or without transarterial embolization) in patients with head, neck, and spine tumors treated between 2012-2023. Baseline demographics, angiographic imaging, tumoral pathology, and relevant clinical variables were retrospectively reviewed and descriptively analyzed.

**Result:** A total of 55 patients underwent direct percutaneous embolization (19 of whom also received transarterial embolization either before [n=7] or after [n=12] DPE). The most commonly embolized lesions were malignant carcinomas (n=16), followed by juvenile nasopharyngeal angiofibromas (n=11), paragangliomas (n=8), hemangiomas (n=7), and others (hemangioblastoma, schwannoma, neurofibroma, esthesioneuroblastoma, meningioma and hemangiopericytoma). The most common locations were sinonasal/nasopharyngeal (n=21), followed by scalp/subcutaneous (n=18), carotid body (n=6), spinal/paraspinal (n=4), skull base (n=5), and intracranial (n=1). A median of 4 (interquartile range 4-6) twenty-two-gauge spinal needles were used per embolization with a median fluoroscopy time of 50.5 (23.2-77.8) minutes resulting in median radiation exposure of 3055 (840.5-5053.5) mGy. Seven patients received more than one embolic agent, with *n-*butyl cyanoacrylate (glue, n=44, 81.5%) being the mostly commonly used embolic, followed by ethylene-vinyl alcohol copolymer (Onyx, n=7, 12.9%), 98% dehydrated ethanol (n=7, 12.9%), sodium tetradecyl sulfate (n=2, 3.7%) and poly-vinyl alcohol particles (n=1, 1.8%). The median volume of embolic agent injected was 5.5(4-7.6) mL resulting in total/near-total (90%-99%) angiographic devascularization in 74.5% cases. The median operative blood-loss was 250(75-700) mL. One patient underwent trans-calavarial DPE for a cerebellar hemangioblastoma and suffered diffuse subarachnoid hemorrhage from profuse tumoral bleeding. One patient had an asymptomatic parent-vessel occlusion from retrograde embolic extension.

**Conclusion:** Our single-center study reinforces prior experience that DPE of Sino-nasal carcinomas, angiofibromas and paragangliomas with adhesive and non-adhesive liquid embolic agents is safe, feasible and effective. Further, it suggests that these benefits may also be extended to non-traditional head, neck and spine tumors. Caution must be exercised when applying these techniques to intracranial tumors with robust intratumoral arteriovenous shunting.

## Introduction

Trans-arterial embolization (TAE) in the management of hypervascular tumors of the head, neck, and spine has been shown to be a safe and effective adjuvant to surgical resection or palliation.^1^ However, the tortuosity of parasitized vasculature and risk of embolic ischemic stroke may preclude its use.^1,2^ Since the introduction of direct percutaneous embolization (DPE) via intra-tumoral injection of embolic material in the 1990’s, several case-reports and short-series have supported the safety and efficacy of this technique.^1,3^ In the absence of level A/B evidence from randomized trials to support use of TAE/DPE for tumor embolization,^1,3^ we provide further retrospective data favoring DPE of head, neck, and spine tumors using liquid embolic agents (*ethylene-vinyl alcohol copolymer, Onyx;™* or *n-butyl cyanoacrylate, n*-BCA; *sodium tetradecyl sulfate*, Sotradecol;^®^ or *polyvinyl-alcohol* particles) in 55 consecutive patients at a tertiary-referral University Hospital. Further, we add to the contemporary practice of trans-calavarial embolization of infratentorial tumors reported in three patients prior, ^4,5^ by reporting our experience with DPE of a cerebellar hemangioblastoma. We describe the technical aspects of the procedure and its efficacy in reducing intraoperative blood loss.

## Methods

This was a retrospective, observational, study conducted at our institution from a prospectively maintained database after receiving a waiver from the Institutional Review Board on the need to obtain informed patient consent given the retrospective nature of the study. The study was performed under the guidelines outlined by the Declaration of Helsinki. Patients with hypervascular lesions of head, neck, or spine that were referred to the neurointerventional service for consideration of embolization between 2012 and 2023 at The University of Minnesota were retrospectively reviewed. All patients meeting inclusion criteria underwent DPE with or without TAE, and had adequate clinico-radiographic data including their operative and pathology reports available for review. Patients undergoing TAE and/or DPE received general anesthesia with or without neuromonitoring (electroencephalography, visual evoke potentials, and somatosensory-evoked potentials). All patients received at least one transfemoral or transradial diagnostic angiogram to delineate the arterial and venous angioarchitecture. Depending on the anatomic location of the tumor, the following craniocervical arteries were injected: internal carotid, external carotid, vertebral, subclavian, costocervical, and thyrocervical. The following angiographic/radiographic findings were factored into favouring DPE over TAE: [1]*Tumor location:* endonasal or intra-oral or intra-aural; soft-tissue tumor <=5 inches from skin surface; vertebral body tumor with a preserved pedicle [2] *Parasitized arteries*: small caliber (<0.5mm) and/or sever tortuosity precluding superselective catheterization, and presence of >=4 pedicles [3] *Dangerous collaterals* to the intracranial circulation, and [4] *Minimal or sub-total residual* parenchymal opacification after TAE.

### TAE Technique

Superselective catheterization of the parasitized artery was performed to look for dangerous intracranial collaterals and assess reflux potential. Typically, a 1.6F or 1.3F tip microcatheter (Headway™ Duo 156cm and 167cm respectively, *Microvention, Aliso Viejo, CA)* was used; with both the contrast injection and subsequent embolization performed using the same type of syringe (i.e., either a regular 1cc luer-lock syringe or 1cc high-pressure polycarbonate syringe) to ensure a visually comparable reflux assessment between contrast and embolic injections. Per the discretion of the treating neurinterventionalist, a 1.6F tip balloon-microcatheter (Scepter™ Mini, MicroVention™, Aliso Viejo, CA) was used to minimize reflux. The liquid embolic was injected through a 1cc syringe under constant negative roadmap until complete stasis of flow within each feeding pedicle was achieved.

### DPE Technique

The percutaneous site was prepped and draped using standard sterile technique prior to obtaining arterial access. A prophylactic antibiotic (e.g., Cefazolin) was administered 30-minutes before DPE. Once baseline angiography was completed, the diagnostic catheter was parked in the parent vessel (e.g., ipsilateral external carotid artery for patients with juvenile nasoangiofibromas). Approximately 80 international units (IU) per kilogram of unfractionated Heparin was administered intravenously, with an additional 1000 IU administered every 60-minutes thereafter. At least three 22-gauge Quincke-tip spinal needles (3.5” or 5” long) were prepped by removing their stylet’s and attaching a 5” Microbore Extension Set. Under live roadmap guidance, a prepped 22-gauge spinal-needle was advanced into the tumor, with blood-return suggestive of penetration into the tumor-bed. Next, a ‘*parenchymogram’* was performed by injecting contrast into the tumor using a 3-cc syringe (Figure: 1). Maintaining anionic precautions, the dead space of the needle and Microbore tubing was then primed with DMSO or 5% Dextrose using a 1-cc syringe, followed by injection of Onyx or n-BCA respectively under blank roadmap guidance. While exchanging for the embolic syringe, the Luer lock extension tubing was occluded to avoid reflux of blood, which may otherwise result in precipitation of embolic within the needle. The duration of the injections varied from a few seconds to several minutes. If the embolic appeared to enter a vein or an arterial pedicle, we stopped the injection and waited ∼30 seconds to 1 minute. This was repeated until the embolic was able to penetrate the desired segment of the tumor. The injection was stopped if there was significant retrograde filling of an arterial pedicle, or a venous outflow vessel, or dangerous intracranial anastomoses. Intermittent angiography was performed to determine the extent of devascularization and to target the next neovascular compartment. At the end of each embolization, the Microbore tubing was disconnected and the stylet reintroduced prior to removal of the needle. If the embolic material stopped advancing or if there was intratumoral interstitial extravasation, multiple needles were used to maximize embolization within the different tumoral compartments.

**Figure 1:**
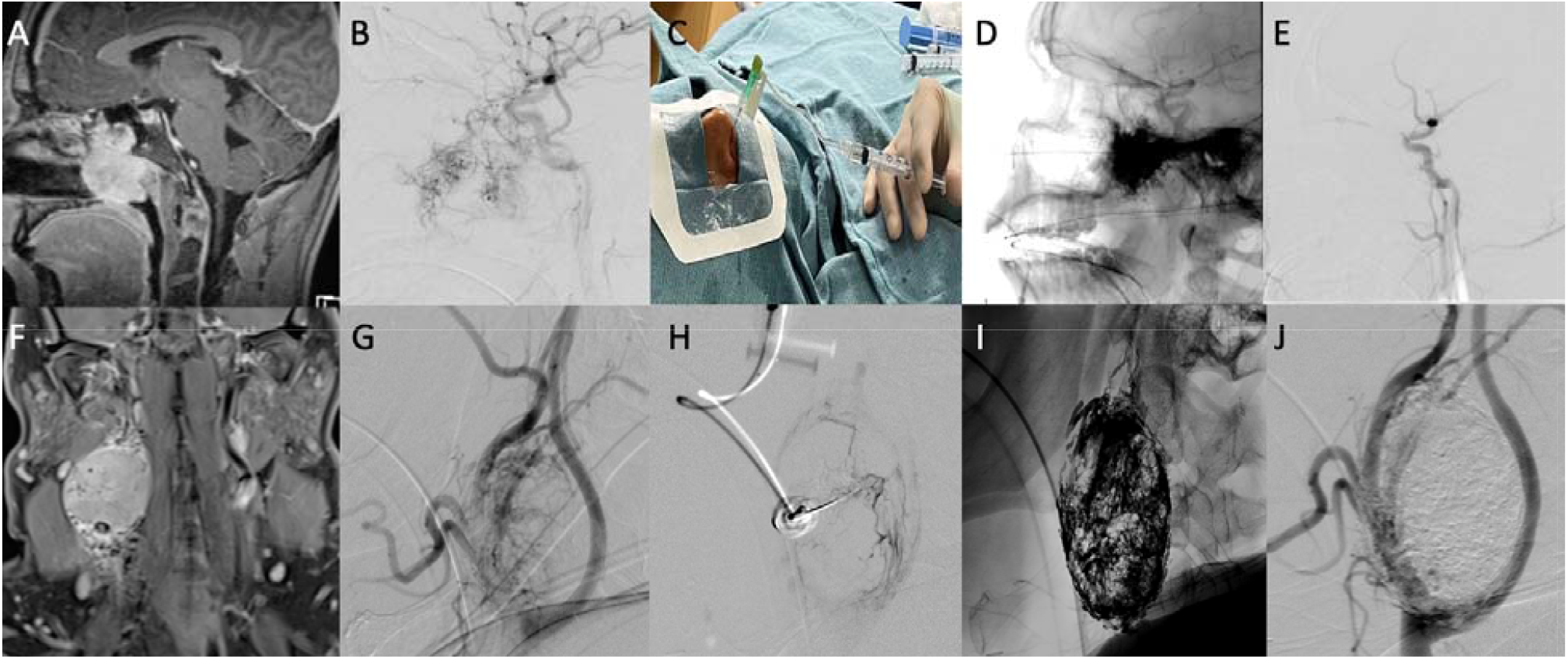
Direct percutaneous embolization Technique. Post-gadolinium MR imaging demonstrates pre-embolization morphology of a juvenile nasoangiofibroma (JNA, A) and a carotid-body paraganglioma (CBP, F). Baseline trans-femoral angiography in patient with the JNA identified several parasitized arterial feeders arising from the ipsilateral internal maxillary, middle meningeal, vidian and inferolateral trunk branches (B); while the patient with CBP has parasatization of the ipsilateral ascending pharyngeal and occipital arteries (G). A 22-gauge spinal-needle attached to a 5-inch Microbore tub-extension was advanced directly into the tumor under direct visual and bi-planar roadmap guidance, followed by injection of contrast using a 3cc syringe to obtain a *parenchymogram* (C, H). The needle is then primed with 5% dextrose, followed by 25% n-BCA injection under blank roadmap guidance forming a dense embolic cast (D, I). Final angiography demonstrates total and near-total reduction in tumor-blush of the JNA and CBP respectively (E, J)

With either technique, the choice of the liquid embolic agent was left to the discretion of the treating neurointerventionalist. The degree of residual parenchymal staining of the tumor on biplane angiography at the end of the treatment was graded by two neurointerventionalists as being: total (99%-100%), near-total (90%-98%), sub-total (50%-89%) or minimal (<50%). Total or near-total obliteration was considered a technical success.

## Results

A total of 55 patients underwent direct percutaneous embolization. There were 37 (67.3%) males, with a median age of 46.5 (27-65) years. Approximately, 14 (25.5%) patients had a medical history of hypertension and 5 (9.1%) had diabetes mellitus. The most common locations for DPE were endonasal/nasopharynx (n=21), followed by scalp/subcutaneous tissue (n=18), carotid body (n=6), spinal/paraspinal (n=4), skull base (n=5), and intracranial (n=1). The most commonly embolized lesions were malignant carcinomas (n=16), followed by juvenile nasopharyngeal angiofibromas (n=11), paragangliomas (n=8), and hemangiomas (n=7). The most common malignant carcinoma was spinal metastatic disease from renal cell carcinoma (n=5), with papillary thyroid carcinoma (n=2), adenocarcinoma (n=2), squamous cell carcinoma (n=2) and sarcoma (n=2) amongst others (Figure: 2). A median of 4 (interquartile range 4-6) needles were used per embolization with a median fluoroscopy time of 50.5 (23.2-77.8) minutes. The median radiation dose per patient was 3055 (840.5-5053.5) mGy. Seven patients received DPE with more than one embolic agent. The most commonly used agent was *n-*butyl cyanoacrylate (glue, n=44, 81.5%), followed by ethylene-vinyl alcohol copolymer (Onyx, n=7, 12.9%), 98% dehydrated ethanol (n=7, 12.9%), sodium tetradecyl sulfate (n=2, 3.7%) and poly-vinyl alcohol particles (n=1, 1.8%). Nineteen of these 55 patients also received transarterial embolization, with 12 (63%) of those performed after DPE. A median of 2 parasitized pedicles were embolized trans-arterially with a Headway Duo microcatheter being the most commonly chosen microcatheter (n=12), followed by Excelsior SL 10 (Target Therapeutics, Boston Scientific) microcatheter (n=5), and Scepter Mini dual-lumen balloon (Microvention, Aliso Viejo, CA) microcatheter (n=2). The median volume of embolic agent injected was 5.5(4-7.6) mL per patient. Total (99%-100%), near-total (90%-98%) and sub-total (50%-89%) reduction in angiographic tumoral-blush was obtained in 20%, 54.5%, and 25.5% cases respectively. Fifty-one patients (93%) achieved atleast 80% reduction in tumoral vascularity. The technical success rate was highest in patients with juvenile nasoangiofibroma (n=11, 100%), followed by schwannoma (n=2, 100%), paraganglioma (n=7, 88%), neurofibroma (n=4, 75%), malignant carcinoma (n=16, 62.5%), and hemangioma (n=7, 57.1%). Of the 55 patients receiving DPE, 50 were followed by surgical resection, with three patients treated palliatively for refractory blood-loss anemia, and two undergoing treatments for localized pain. The median interval between embolization and surgical resection was 1 (1-4) day. The median operative blood-loss was 250(75-700) mL, with 12 patients (22.2%) receiving a postoperative blood transfusion. Only one of the 55 patients undergoing DPE had to be taken for emergent tumor resection due to uncontrolled tumoral hemorrhage. This patient had a large recurrent cerebellar hemangioblastoma which was embolized by advancing a 22-gauge spinal-needle directly through a previously placed sub-occipital titanium mesh (Figure: 3). The other complication from DPE included retrograde extension of n-BCA into parasitized vertebral artery feeders resulting in a clinically asymptomatic vertebral artery occlusion (Figure: 4). While one patient with a Jugular paraganglioma suffered a peripheral cranial-nerve VII palsy; however, this was likely due TAE in which Onyx-18 was injected under proximal balloon-occlusion in the posterior auricular artery resulting in devascularization of the trans-mastoid segment of the seventh cranial nerve without any identifiable drop-out in electromyographic activity during the embolization.

**Figure 2:**
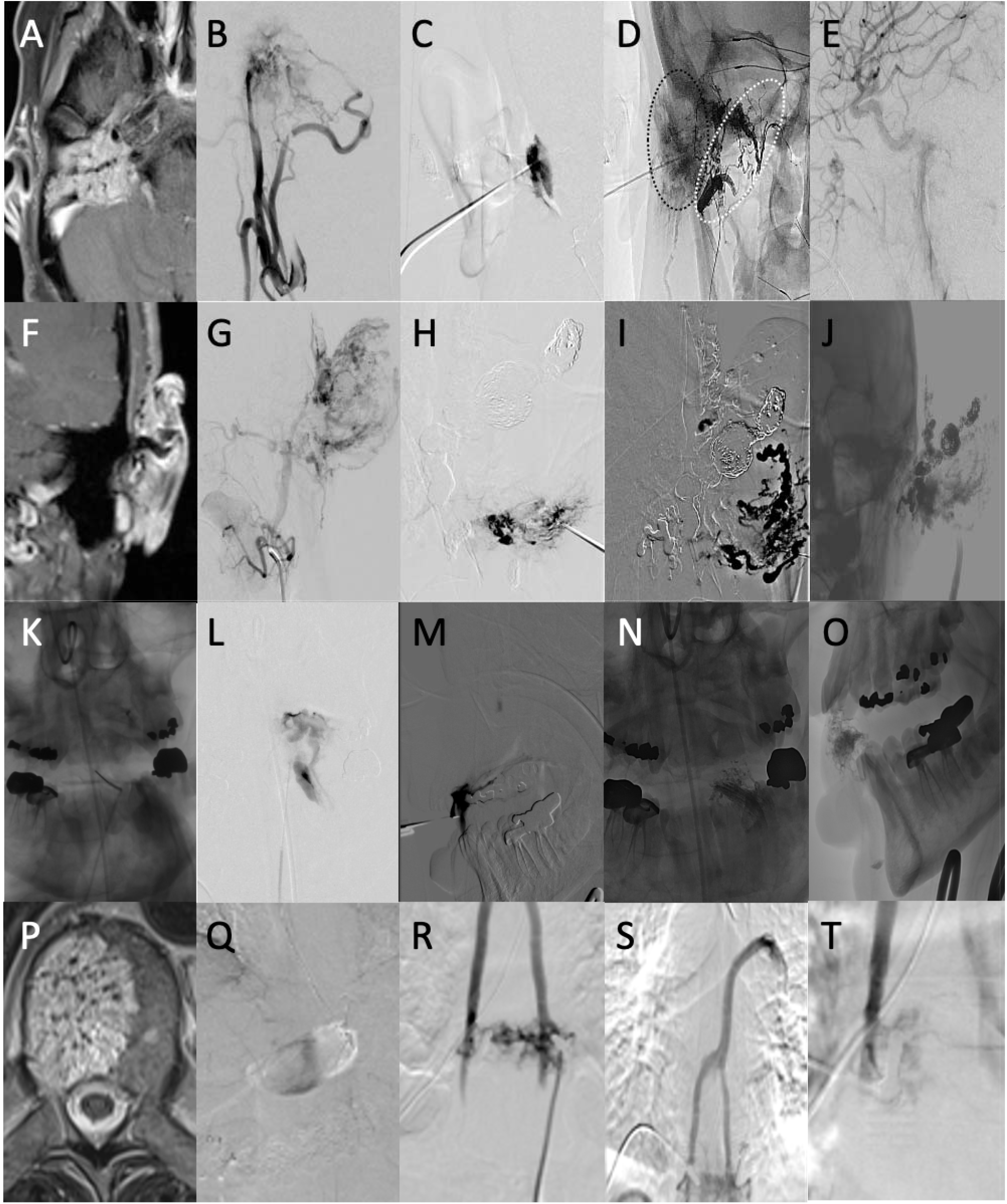
Manifold application of Direct percutaneous embolization techniques. ***Trans-aural:*** Post-gadolinium axial brain MRI demonstrates a jugular paraganglioma (A) eroding through the tympanic membrane into the external auditory canal. Baseline angiography identified extensive parasatization of the right ascending pharyngeal, posterior auricular, and occipital arteries (B) which were selectively catheterized with a Scepter-Mini microcatheter for Onyx-18 embolization reducing tumoral vascularity by ∼80%. The tumor was now directly punctured with a 22-gauge spinal needle advanced into the external auditory canal under direct visualization. Parenchymography (C) was followed by 25% n-BCA embolization (D), resulting in near-total obliteration of tumoral vascularity. Note that DPE achieved a more diffuse penetration of n-BCA within the capillary-bed *(black-dotted oval)* when compared to the trans-arterial Onyx-cast, which was essentially restricted to the larger *(white-dotted oval)* proximal vasculature (D). The tumor was resected the next day with an estimated blood loss of 300mL. ***Per-aural:*** Post-gadolinium coronal brain MR imaging demonstrates a complex left auricular arteriovenous malformation (F) that had been previously treated at an outside facility 2-years prior with trans-arterial coiling and liquid embolic embolization. Patient now presented with recurrent bleeding over 3-months. Trans-femoral angiography demonstrated a diffuse nidus (∼4.4cm x 7.4cm) fed by parasitized branches of the left superficial temporal, middle meningeal, and posterior auricular arteries (G). With the patient intubated and maintained under optimal analgesia, parenchymography was performed by directly puncturing the helix and lobule of the left ear (H). This was followed by DPE with 25% n-BCA, as well as, dehydrated ethanol (5cc of 98% dehydrated ethanol with 1cc of lipiodol) under roadmap guidance (I), resulting in near-total obliteration of left ear vascularity (J). This was immediately followed by auriculectomy and aural reconstruction with an estimated blood-loss of 50 mL. ***Trans-oral***: A patient in his mid-50’s with a previously resected left lingual hemangioma presented with 2-months of recurrent bleeding. Under direct visual guidance a 22-gauge spinal-needle was inserted into the hemangioma (K) to perform parenchymography (L), followed by 25% n-BCA embolization under roadmap guidance (M). The embolic cast (N, O) was resected the next day with an estimated blood-loss of 10 mL. ***Trans-pedicular:*** Post-gadolinium axial MRI of the thoracic spine identified a T10 vertebral body hemangioma in a man with back-pain with focal tenderness (P). Spinal angiography identified dense blush over the T10 vertebral body from the left and right T10 segmental arteries (Q). With one Jamshidi needle each advanced trans-pedicularly into the T10 vertebral body, parenchymography was performed revealing capillary blush of the hemangioma followed by venous outflow through the azygos system into the right atrium (R, S). Dehydrated ethanol (98%) was injected keeping a column of contrast proximal and distal to the alcohol with an intervening air bubble between the columns of contrast and ethanol for better visualization, as well as, to avoid dilution of ethanol with contrast. A total of 6cc of ethanol was injected resulting in approximately 60% reduction in contrast blush of the hemangioma.

**Figure 3:**
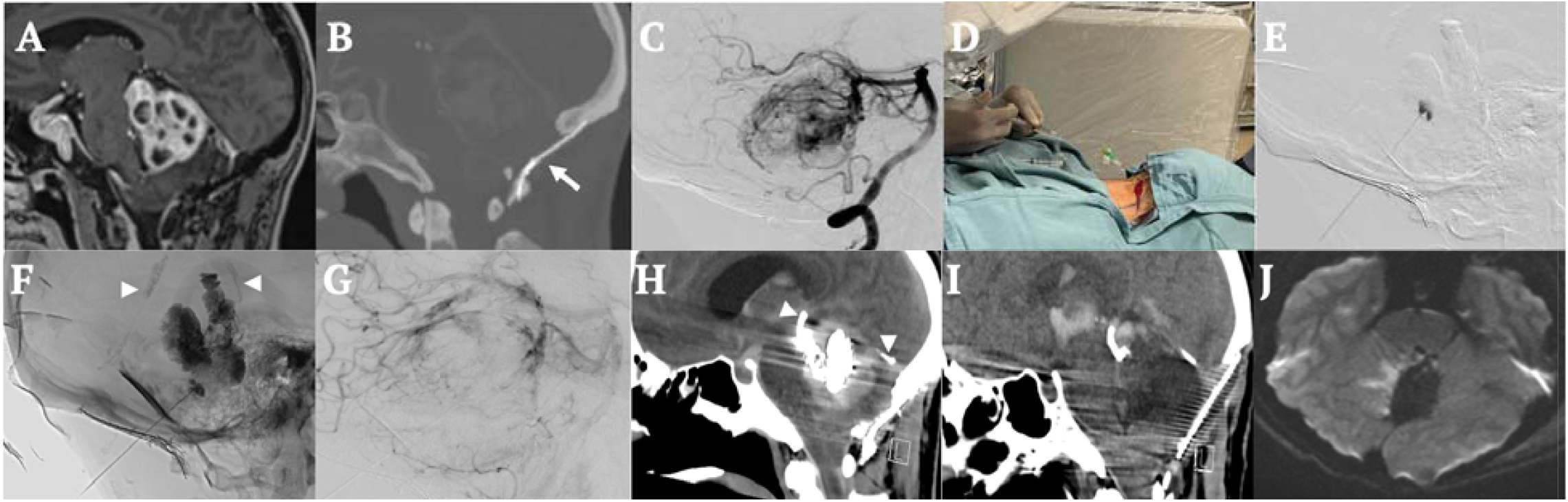
Intracranial Trans-calavarial DPE. A young man in his forties with past medical history of a partially-resected cerebellar hemangioblastoma was found to have tumoral re-growth after he presented with new-onset headache and projectile vomiting. Neurological examination identified bilateral cranial-nerve VI palsy. Post-gadolinium MR imaging of the brain (a) and CT angiography (b) demonstrated an avidly enhancing, cystic lesion with parasitized feeders from bilateral superior-cerebellar arteries and the left posterior-inferior cerebellar artery (c). There was extensive arteriovenous shunting through the tumor with venous drainage through the superior vermian vein into the vein of Galen. Considering the potential for incomplete embolization with TAE and the inherent risk of ischemic stroke, we decided to pursue DPE by advancing a 22-gauge spinal needle through the previously placed titanium mesh under roadmap guidance (d). ‘Parenchymography’ (e) was performed, followed by injection of 25% n-BCA forming a dense embolic cast (f). After 4 separate needle insertions, we were able to achieve approximately 60% reduction in the tumoral vascularity with residual blush over the postero– superior tumor (g). Immediate post-procedural head CT demonstrated peri-tumoral hemorrhage with extension into the subarachnoid and intraventricular space resulting in obstructive hydrocephalus (h). An emergent external ventricular drain was placed, followed by immediate suboccipital craniectomy and tumor resection. On direct visualization, the non-embolized tumor was noted to be hemorrhaging profusely causing mass-effect resulting in diffuse cerebellar venous congestion. The embolized portion of the tumor was resected *en-bloc*, with intra-procedural EBL ∼1000mL. He was extubated on post-operative day#3 after a reassuring MRI scan (i). Six-month clinic follow-up demonstrated mild left upper and left lower extremity weakness with improvement in bilateral abducens nerve paresis.

**Figure 4:**
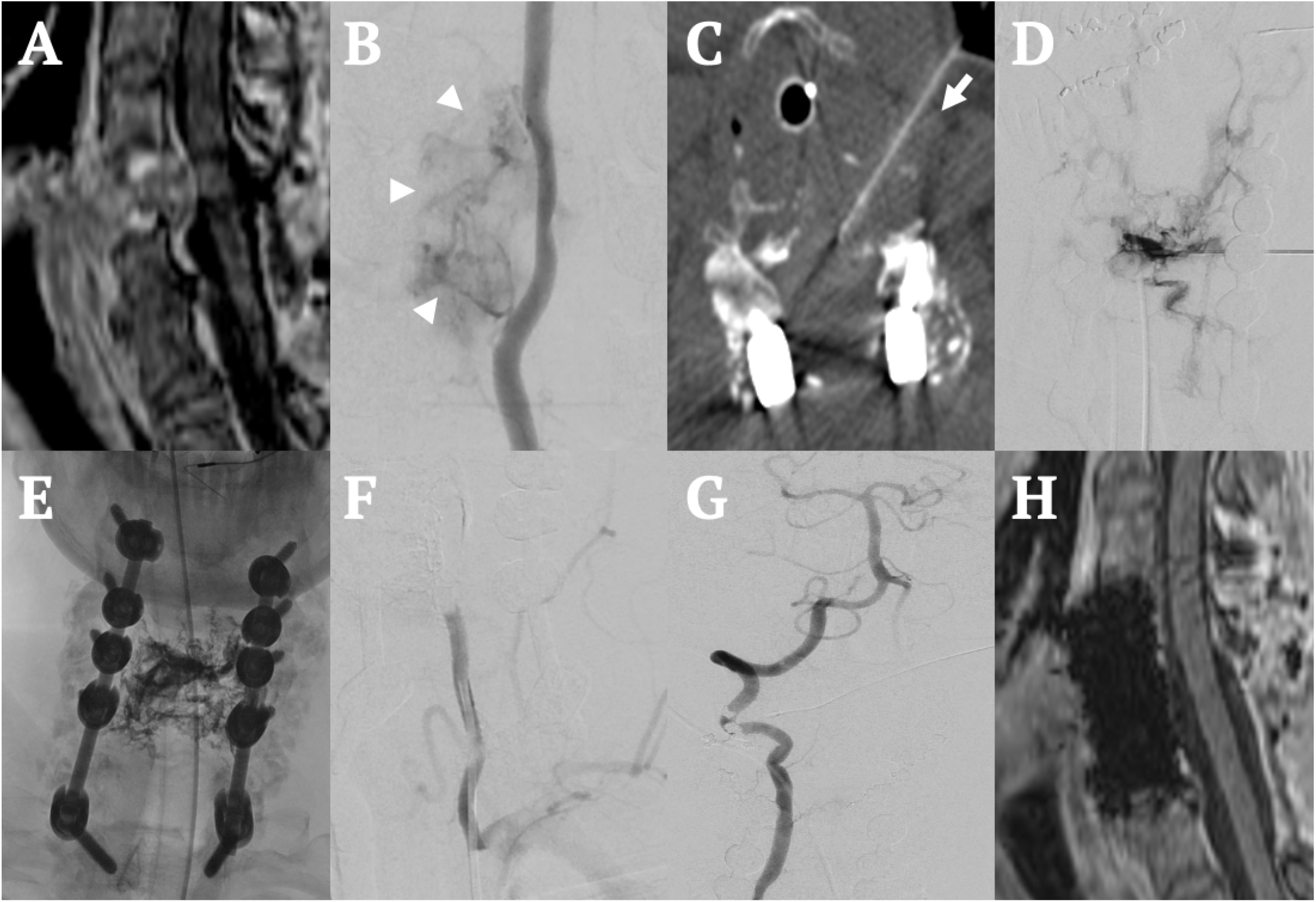
Parent vessel-occlusion from post-capillary retrograde reflux of liquid-embolic. A middle-aged woman with a pathologic fracture of C5 vertebrae from metastatic renal cell carcinoma status post C3-T1 spinal fusion surgery two-weeks ago, presented with progressive left greater than right upper extremity weakness. She was found to have spinal-cord compression on post-gadolinium T1-weighted sagittal MRI (a). The vertebral arteries were co-dominant with parasitized feeders from left greater than right V2 segments supplying the vertebral body metastases (b). Considering the risk of vertebral artery ischemic stroke with TAE and the lack of a trans-pedicular approach from prior instrumentation, we performed DPE by advancing a 21-gauge spinal-needle under CT guidance (c). ‘Parenchymography’ (d) was followed by 33% n-BCA injection forming a dense embolic cast (e). Unfortunately, despite a cautious injection of liquid embolic, there was retrograde penetration of embolic material via the parasitized arterial branches into the left vertebral artery resulting in its occlusion (f). Subsequent right vertebral artery angiography did not show any intracranial vessel occlusions or capillary phase defects (g). The patient’s neurological exam remained unchanged and she underwent a C5 corpectomy with C4-C6 anterior cervical fusion with an estimated blood loss of 20cc (h).

## Discussion

In this series, which is the largest to-date describing the use of percutaneous embolization for head, neck and spine lesions, we find it to be safe (1.8% major complication rate) and effective (93% achieving 80% or higher angiographic devascularization). Further, this is the largest series to report DPE experience with n-BCA (used in over 80% cases) via a myriad of approaches (trans-cutaneous, trans-mucosal, trans-aural, per-aural, per-oral, trans-pedicular and trans-calavarial). Finally, we add to the currently limited experience of trans-calavarial DPE and summarize the relevant literature.

It is now well-established that preoperative trans-arterial embolization of hypervascular head, neck, and spine tumors reduces operative times and blood-loss.^1^ Several agents have been utilized over the years, driven largely by operator preference, with liquid embolic agents (e.g., Onyx and n-BCA) being increasingly utilized over particles given the risk of injury to vasa-nervosum and delayed recanalization.^1,6^ However, parasitized arterial feeders can occasionally be very tortuous making catheterization difficult. Some tumors may also have a high degree of arterio-venous shunting, further obscuring its arterial anatomy.^2,8^ In addition, dangerous external-carotid to internal-carotid or vertebral artery anastomoses may develop in tumor-beds making trans-arterial embolization high-risk.^1,2^ To bypass these limitations, direct percutaneous embolization was initially introduced in 1994 as an alternative approach to devascularize tumors,^2,7^ with several case-series since providing retrospective data on the safety and efficacy of this technique.^6-11^ Interestingly, this early experience with DPE has also shown that it results in greater penetration into the capillary-bed of these tumors when compared to TAE, wherein occlusion of the proximal feeders may result in a high-degree of angiographic devascularization without any significant reduction in operative blood-loss as the capillary-bed may remain ‘alive’ via collateral pathways (Figure 2, D).^1,8^ Thus, TAE may not always be achievable, and even when it is feasible, its use alone may result in radiographic devascularization without conferring a net clinical benefit .^1,8^

Several embolic agents have been used for DPE including Onyx, n-BCA, SQUID-12, and Polyvinyl alcohol or trisacryl gelatin microspheres, with particles found to be associated with a higher risk of cranial neuropathies from devascularization of vasa-nervosum.^1^ In our series, we predominantly (81.5% cases) used n-BCA diluted to a 25% concentration by diluting it with Lipiodol (i.e., 0.9mL of n-BCA diluted in 2.7mL of Lipiodol). While we did use Onyx-18 in ∼13% cases, we found that low-concentration n-BCA allowed for a more centripetal distribution of embolic material within the tumor-bed. Further, lower concentrations of n-BCA have higher viscosity allowing for more controlled and pronged injections. In addition, it’s almost immediate polymerization could become useful in cases of extravasation.^5,6^ Despite its rapid polymerization, we did not find any significant increase in the rate of needle-occlusion with n-BCA when compared to Onyx-18.

The radiographic goal of pre-operative embolization is generally considered to be atleast 80% devascularization on final angiography,^1^ which was achieved in 93% cases in our series with ∼75% achieving total (99%-100%)/near-total (90%-98%) angiographic devascularization. This is comparable to other single institutional studies that have shown similar rates of devascularization with DPE of head and neck tumors,^1-3,8-11^ vascular malformations,^12^ and spinal tumors.^13^

Intra-operative needle puncture and embolization of intracranial tumors at the time of their resection has been well described.^14,15^ However, only a handful of case-reports have described transcalavarial puncture for DPE of intracranial tumors (Table 1). Seven of the eight previously reported cases of transcalavrial DPE have utilized prior craniectomies from aborted resection attempts ^3-5,16,17^ or pathologic bony erosion;^5^ while one patient with an infratentorial endolymphatic-sac tumor underwent a retroauricular craniotomy to facilitate DPE with subsequent tumor resection via the same craniotomy.^5^ Among the three reported cases of transcalavrial DPE performed for infratentorial tumors,^4,5^ one patient had a cerebellar hemangioblastoma while the other two patients had petro-temporal endolympatic-sac tumors; undergoing Onyx-34, 15% n-BCA and 15% n-BCA embolization resulting in 50%, near-total (>90%) and total tumoral devascularization respectively. In our series, we treated one patient with a recurrent cerebellar hemangioblastoma with DPE by advancing a 22-gauge spinal-needle (under bi-planar fluoroscopic/roadmap guidance) via a previously placed titanium mesh (Figure 3). We used 25% n-BCA resulting in nearly 60% devascularization after four punctures. Similar to our patient (Figure 3), the only other previously reported case of DPE for a cerebellar hemangioblastoma, was also immediately resected in the operating room where the non-embolized residual was noted to bleed profusely resulting in an estimated blood-loss of 1000mL.^4^ However, unlike the prior case where the tumor was homogenous and solid (type IV hemangioblastoma), our patient had a microcystic tumor with multiple gadolinium-enhancing cysts (type-III hemangioblastoma).^4,18^ Cysts with enhancing walls have been shown to contain neoplastic cells, with dilated and necrotic vascular spaces on histologic examination, and suffering higher intraoperative blood-loss than the most common type (i.e. type II) hemangioblastomas.^18,19^ The authors believe that the high degree of arteriovenous shunting within the neoplastic cystic walls, may have resulted in embolization of n-BCA into the tumor’s primary draining vein (superior vermian) and the straight sinus, turning the residual non-embolized tumor into a functional arteriovenous malformation with its venous outflow obstructed, causing it to bleed immediately post-embolization. This required emergent tumor decompression and resection with an estimated blood-loss of 1000mL.

**Table 1.**
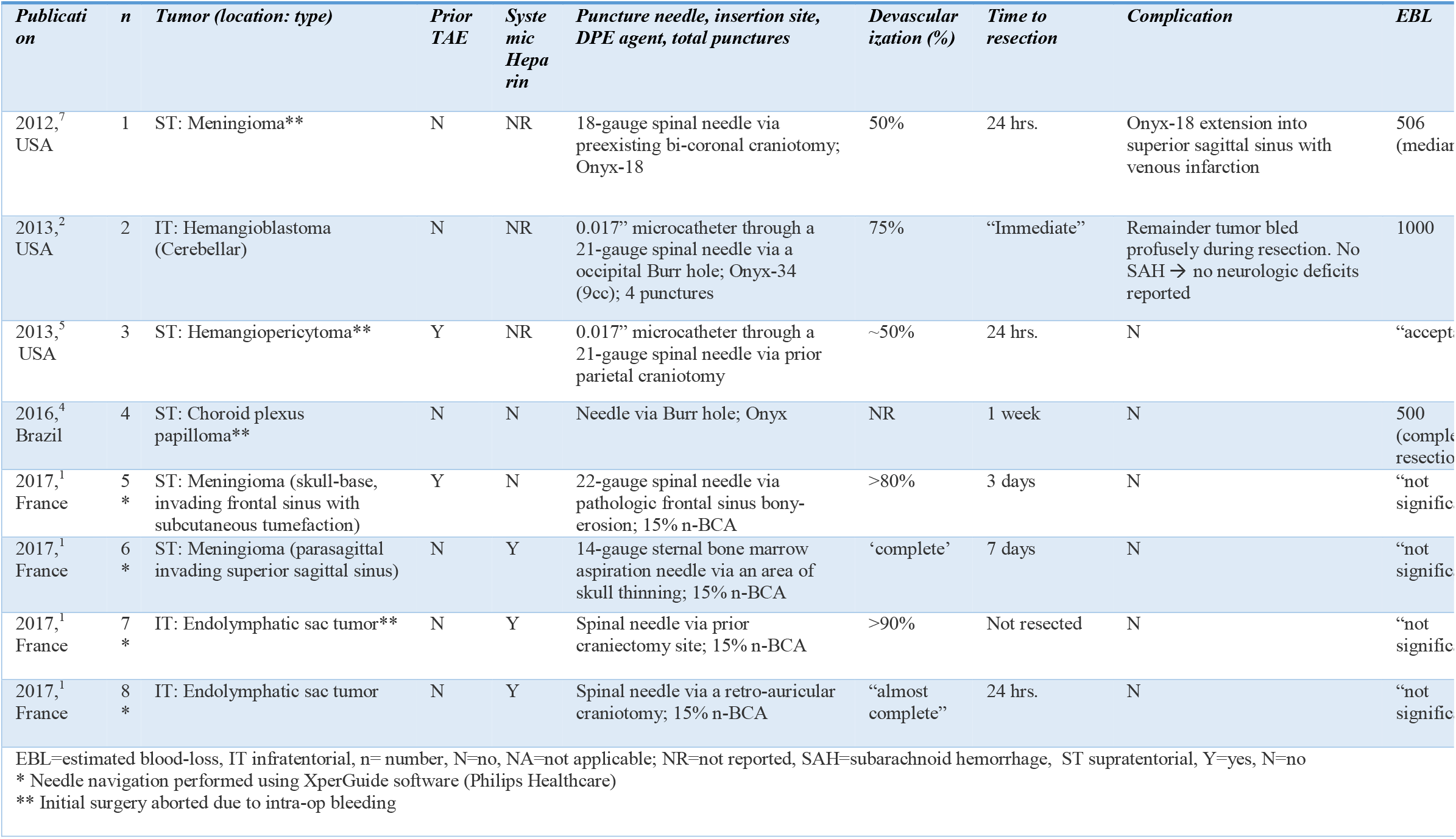
Trans-Calavarial Direct Percutaneous Embolization of Intracranial Tumors.

| *Publication* | *n* | *Tumor (location: type)* | *Prior TAE* | *Systemic Heparin* | *Puncture needle, insertion site, DPE agent, total punctures* | *Devascularization (%)* | *Time to resection* | *Complication* | *EBL* |
| --- | --- | --- | --- | --- | --- | --- | --- | --- | --- |
| 2012,<sup>7</sup> USA | 1 | ST: Meningioma** | N | NR | 18-gauge spinal needle via preexisting bi-coronal craniotomy; Onyx-18 | 50% | 24 hrs. | Onyx-18 extension into superior sagittal sinus with venous infarction | 506 (mediar |
| 2013,<sup>2</sup> USA | 2 | IT: Hemangioblastoma (Cerebellar) | N | NR | 0.017" microcatheter through a 21-gauge spinal needle via a occipital Burr hole; Onyx-34 (9cc); 4 punctures | 75% | "Immediate" | Remainder tumor bled profusely during resection. No SAH → no neurologic deficits reported | 1000 |
| 2013,<sup>5</sup> USA | 3 | ST: Hemangiopericytoma** | Y | NR | 0.017" microcatheter through a 21-gauge spinal needle via prior parietal craniotomy | ~50% | 24 hrs. | N | "accept |
| 2016,<sup>4</sup> Brazil | 4 | ST: Choroid plexus papilloma** | N | N | Needle via Burr hole; Onyx | NR | 1 week | N | 500 (comple resectio |
| 2017,<sup>1</sup> France | 5 * | ST: Meningioma (skull-base, invading frontal sinus with subcutaneous tumefaction) | Y | N | 22-gauge spinal needle via pathologic frontal sinus bony-erosion; 15% n-BCA | >80% | 3 days | N | "not signific |
| 2017,<sup>1</sup> France | 6 * | ST: Meningioma (parasagittal invading superior sagittal sinus) | N | Y | 14-gauge sternal bone marrow aspiration needle via an area of skull thinning; 15% n-BCA | 'complete' | 7 days | N | "not signific |
| 2017,<sup>1</sup> France | 7 * | IT: Endolymphatic sac tumor** | N | Y | Spinal needle via prior craniectomy site; 15% n-BCA | >90% | Not resected | N | "not signific |
| 2017,<sup>1</sup> France | 8 * | IT: Endolymphatic sac tumor | N | Y | Spinal needle via a retro-auricular craniotomy; 15% n-BCA | "almost complete" | 24 hrs. | N | "not signific |
| EBL=estimated blood-loss, IT infratentorial, n= number, N=no, NA=not applicable; NR=not reported, SAH=subarachnoid hemorrhage, ST supratentorial, Y=yes, N=no<br>* Needle navigation performed using XperGuide software (Philips Healthcare)<br>** Initial surgery aborted due to intra-op bleeding |  |  |  |  |  |  |  |  |  |

The other DPE related complication encountered in our series was immediate retrograde extension of 25%n-BCA via the parasitized arterial feeders into the left vertebral artery in a patient with co-dominant vertebro-basilar circulation (Figure 4). The patient remained clinically asymptomatic. However, post-capillary retrograde extension of embolic agents has been reported to result in significant clinical impairments, usually immediately post-embolization while the glue is still in a liquid state.^11^ However, delayed embolization thought to have been caused by fragmented, polymerized glue has also been reported.^20^ While some have used compliant balloons within the parent vessel (e.g., internal carotid or vertebral arteries) during DPE for this particular reason; the recurrent inflation of these intravascular polyurethane balloons may also confer an additive thromboembolic risk, with some reporting migration of embolic agents despite balloon-inflation.^20^

## Conclusion

Direct percutaneous embolization of head, neck, and spine tumors is safe and effective. Our single-center study reinforces prior experience that DPE of sino-nasal carcinomas, angiofibromas and paragangliomas with adhesive and non-adhesive liquid embolic agents achieves good penetration of capillaries within the tumor-bed significantly reducing operative blood-loss. Further, it suggests that these benefits may also be extended to non-traditional head, neck and spine lesions such as schwannomas, neurofibromas, and hemangiomas. Caution must be exercised when applying these techniques to intracranial tumors with high intratumoral arteriovenous shunting.

## Data Availability

All data produced in the present study are available upon reasonable request to the authors

## Acknowledgements

None

## Abbreviations

CTA: computed tomography angiogram
JNA: juvenile nasopharyngeal angiofibroma
n-BCA: n-butyl cyanacrylate
MCA: middle cerebral artery
TAE: Trans-arterial embolization

## Notes

**Conflict of Interests:** B.D.J is a consultant for *Microvention, Medtronic* and *Stryker*. On behalf of all other authors, the corresponding author declares that there are no competing conflicts of interest.

### Competing Interest Statement

B.D.J is a consultant for Microvention, Medtronic and Stryker. On behalf of all other authors, the corresponding author declares that there are no competing conflicts of interest.

### Funding Statement

This study did not receive any funding

### Author Declarations

This was a retrospective, observational, study conducted at our institution from a prospectively maintained database after receiving a waiver from the Institutional Review Board on the need to obtain informed patient consent given the retrospective nature of the study. IRB of the University of Minnesota waived ethical approval for this work.

